# COVID-19 outbreak rates and infection attack rates associated with the workplace: a descriptive epidemiological study

**DOI:** 10.1101/2021.05.06.21256757

**Authors:** Yiqun Chen, Timothy Aldridge, UK COVID-19 National Core Studies Consortium, Claire F. Ferraro, Fu-Meng Khaw

**Affiliations:** Health and Safety Executive, UK; National Infection Service, Public Health England, UK; National Institute of Health Research Health Protection Research Unit in Behavioural Science and Evaluation at the University of Bristol; Public Health England, UK

**Keywords:** SARS-Cov-2, COVID-19, coronavirus, transmission, outbreak, cluster, outbreak rate, attack rate, settings, workplace, geographical, England

## Abstract

**Background:** A large number of COVID-19 outbreaks/clusters have been reported in a variety of workplace settings since the start of the pandemic. However, information on the rate of outbreak occurrences which helps to identify the type of workplaces that are more likely to experience an outbreak, or infection attack rates which estimates the potential extent of the virus transmission in an outbreak, has not yet been available to inform intervention strategies to limit transmission.

**Objectives:** To link datasets on workplace settings and COVID-19 workplace outbreaks in England in order to: identify the geographical areas and workplace sectors with a high rate of outbreaks; and compare infection attack rates by workplace size and sector.

**Methods:** We analysed Public Health England (PHE) HPZone data on COVID-19 outbreaks in workplaces, covering the time period of 18 May – 12 October 2020. The workplaces analysed excluded care homes, hospitals and educational settings. We calculated the workplace outbreak rates by nine English regions, 151 Upper Tier Local Authorities (UTLAs) and twelve industrial sectors, using National Population Database (NPD) data extracted in May 2019 on the total number of the relevant workplaces as the denominator. We also calculated the infection attack rates by enterprise size (small, medium, large) and industrial sector, using PHE Situations of Interest (SOI) data on the number of test-confirmed COVID-19 cases in a workplace outbreak as the numerator, and using NPD data on the number employed in that workplace as the denominator.

**Results:** In total, 1,317 confirmed workplace outbreaks were identified from HPZone data, of which 1,305 were available for estimation of outbreak rates. The average outbreak rate was 66 per 100,000 workplaces. Of the nine geographical regions in England, the North West had the highest workplace outbreak rate (155/100,000 workplaces), based on 351 outbreaks. Of the UTLAs, the highest workplace outbreak rate was Blackburn with Darwen (387/100,000 workplaces). The industrial sector with the highest workplace outbreak rate was manufacturers and packers of food (1,672/100,000), based on 117 outbreaks: this was consistent across seven of the regions. In addition, high outbreak rates in warehouses were observed in the East Midlands and the North West.

In total, 390 outbreaks were identified from SOI data and 264 of them allowed for estimation of attack rates. The overall median attack rate was 3.4% of the employed persons with confirmed COVID-19 at a workplace with an outbreak. Most of these outbreaks (162) had an attack rate less than 6%. However, in a small number of outbreaks (57) the attack rate was over 15%. The attack rates increased as the size of the enterprise decreased. The highest attack rate was for outbreaks in close contact services (median 16.5%), which was followed by outbreaks in restaurants and catering (median 10.2%), and in manufacturers and packers of non-food products (median 6.7%).

**Conclusions:** Our linked dataset analysis approach allows early identification of geographical regions and industrial sectors with higher rates of COVID-19 workplace outbreaks as well as estimation of attack rates by enterprise size and sector. This can be used to inform interventions to limit transmission of the virus. Our approach to analysing the workplace outbreak data can also be applied to calculation of outbreak rates and attack rates in other types of settings such as care homes, hospitals and educational settings.

## Introduction

Severe acute respiratory syndrome coronavirus 2 (SARS-CoV-2) is a highly transmissible novel virus that has caused a pandemic of the “coronavirus disease 2019” (COVID-19)^1^. On 30^th^ January 2020, the World Health Organisation declared COVID-19 as a public health emergency of international concern, and later declared a pandemic on 11^th^ March 2020^2^. COVID-19 is a highly contagious disease and can spread rapidly without effective control measures. Because of the heterogeneity characteristics of the SARS-CoV-2 transmission, COVID-19 cases are appearing in clusters in different settings ^3 4^.

In October 2020, Public Health England reported 503 COVID-19 outbreaks/clusters in workplace settings in the previous 4 weeks. This is compared to 720 in care homes, 853 in education settings and 89 in hospital settings^5^. A survey conducted by the European Centre for Disease Prevention and Control (ECDC) reported a total of 1377 COVID-19 clusters in workplace settings across 13 EU/EEA countries and the UK between March and July 2020. Most clusters were reported in long-term care (591 clusters) and hospital (241 clusters) settings, followed by food packing and processing (153 clusters), non-food manufacturing (77 clusters) and office settings (65 clusters)^6^. However, the total number of settings and the number of people exposed within these settings (i.e. the denominator data) could vary significantly. Without the denominator information to calculate the rate of outbreaks, it is difficult to know which types of settings are more likely to experience an outbreak.

This study aims to analyse the occurrence of COVID-19 outbreaks in workplace settings in England to understand which industrial sectors are more likely to experience an outbreak and to estimate the potential extent of the transmission in these workplace outbreaks. These will guide further research and control measures. However, the design of this study would not allow the investigation of factors potentially contributing to the outbreaks. A separate study is underway to address this^7^.

This study is part of National Core Study on Transmission and Environment^8^. The Health and Safety Executive (HSE) and Public Health England (PHE) worked collaboratively and, with the appropriate data-sharing agreements in place, linked the relevant datasets to calculate the outbreak rates for different workplace settings and the infection attack rates among workers working in these outbreak settings.

## Methods

We analysed Public Health England (PHE) data on COVID-19 outbreaks in the workplace, covering the time period of 18^th^ May – 12^th^ October 2020. We calculated the rates of outbreaks by geographical area and industrial sector, using the total number of the relevant settings in the country as the denominator. The workplace settings here are defined using the categories in PHE’s surveillance system. They include non-residential settings that are not schools or hospitals, as outbreaks in these settings are recorded and analysed separately^9^.

A COVID-19 cluster is defined as two or more test-confirmed cases of COVID-19 among individuals associated with a setting (i.e. a workplace) with onset dates within 14 days, where information about exposure between the confirmed cases is not available or is absent. A COVID-19 outbreak will be a COVID-19 cluster where direct exposure between at least two of the test-confirmed cases can be identified or information on an alternative source of infection outside the setting is absent for the initial identified cases^10^.

We used data from three sources, namely PHE HPZone dataset, PHE Situations of Interest (SOI) dataset and the HSE National Population Database (NPD) to calculate outbreak rates by geographical area (Regional and Upper Tier Local Authority (UTLA) and industrial sector, and attack rates of individual workplace outbreaks by enterprise size and industrial sector. These three data sources are described in more detail in below.

### 1. HPZone dataset

HPZone is a national web-based system for communicable disease control in England and is PHE’s case management system^11^. It was deployed nationally in England in January 2010 to improve the collation of information on health protection incidents and assist in their management. It has direct import of laboratory data, receiving statutory infectious disease notifications and collecting contextual data of management of infectious disease cases and outbreaks, and other non-infectious environmental threats.

During the COVID-19 pandemic, HPZone provides summary-level information about the COVID-19 situations (i.e. outbreaks/clusters) that local Health Protection Teams (HPTs) are responding to. HPTs receive information about suspected or confirmed cases of COVID-19 directly from workplaces or through ‘coincidence reports’ from NHS Test and Trace, where two or more individuals report in the same workplace. Test-confirmed cases are linked to HPZone through the Second Generation Surveillance System (SGSS), which is the national laboratory reporting system used in England to capture routine laboratory data, including data on infectious diseases. The postcode of the workplace is entered manually by HPTs to HPZone if the outbreak/cluster was informed directly by the workplace, or by the NHS Test and Trace contact tracers if the outbreak/cluster was identified through coincidence reports. All suspected outbreaks are further investigated by the HPTs in liaison with local partners.

The HPZone outbreaks/clusters also have an ‘overall assessment’ assigned to them to verify if a situation is a confirmed outbreak or a cluster of COVID-19 by a team of epidemiologists working with the National Surveillance Cell at PHE. This process, which makes use of the descriptive data on HPZone, is done at a snapshot in time on a weekly basis for the previous week’s new situations. However, outbreaks evolve over time. If the information about these outbreaks is not updated, for example to capture the increased number of confirmed cases as the outbreak develops, the data could underestimate the true size of the outbreak or clusters as more data become available over time about these outbreaks.

### 2. Situations of Interest (SOI) dataset

The Situations of Interest (SOI) dataset is a subset of outbreaks from the HPZone dataset that are deemed to be more complex to manage and includes updates on the number of test-confirmed COVID-19 cases as the outbreaks evolve over time. At the time of the data analysis, there was no formal definition of a SOI. It is used operationally to share understanding of significant outbreaks due to their scale, impact and complexity. SOI outbreaks are more likely to be investigated by an outbreak Incident Management Team (IMT). A SOI outbreak will be updated regularly until transmission is controlled and as such provides a dynamic tool to track the total number of confirmed cases for the outbreak.

### 3. National Population Database (NPD)

The National Population Database (NPD) was first developed for HSE in 2004 and is currently maintained by HSE on a biennial cycle. It includes geographically referenced estimates of the Great Britain (GB) population in Geographic Information System (GIS) layers ^12^. The NPD groups the GB population into five themes: Residential, Sensitive (e.g. schools, care homes, hospitals and prisons), Transport, Workplaces and Leisure. The Workplace layer provides information on individual workplaces including the number of employees, industry type (based on Standard Industrial Classifications (SIC)) and a spatial reference (address and postcode). The workplace information is extracted from the Office for National Statistics (ONS) Inter-Departmental Business Register (IDBR)^13^ at enterprise level, with the data used in this analysis extracted in May 2019. This extract included information for 2 million UK businesses. Very small companies (typically self-employment) and not-for-profit organisations are not included in IDBR. Some IDBR records report zero employee numbers^13^.

Outbreak rates and attack rates were calculated as follows:

1. Outbreak Rate = the number of outbreaks in workplace settings / 100,000 workplaces Outbreak rate is defined as the proportion of workplace settings with COVID-19 outbreaks, expressed as the number of outbreaks per 100,000 workplaces (see: 1)). The numerator is the number of confirmed workplace outbreaks identified from HPZone. The denominator is the total number of workplaces identified from the NPD. We calculated outbreak rates by industrial sector and geographical area.
2. Attack Rate = the number of test-confirmed COVID-19 cases in a workplace outbreak setting / 100 employed in that setting An attack rate measures the proportion of persons in an identified population who become infected during an outbreak^14^. It indicates the potential extent of the transmission in an outbreak. It is defined here as the proportion of workers in a particular workplace that become cases of COVID-19 by the end of the outbreak, expressed as a percentage (see: 2)). The numerator is the number of test-confirmed COVID-19 cases in a workplace outbreak obtained from the SOI dataset. The denominator is the number employed in that workplace obtained from the NPD. We calculated the attack rates by geographical area and industrial sector, separately for small (1-49 employees), medium (50-249 employees), and large (250 employees or more) enterprises.

The list of outbreaks/clusters in the HPZone dataset and the SOI dataset are manually categorised into primary, secondary and tertiary contexts. “Workplace” is one of the primary contexts, for which the secondary contexts (categories) and the tertiary contexts (sub-categories) are listed in Table 1. In our analysis of workplace outbreaks, we included all secondary contexts, and these were mapped against the Standardised Industry Classifications (SIC) before matching them to the denominator dataset.

**Table 1.** Public Health England (PHE) classification of workplace settings, July 2020

| **Workplace setting category** | **Sub-category** |
| --- | --- |
| Primary producers | Fruit and vegetable growers, animal and animal producers |
| Manufacturers and packers – food | Abattoir, meat products, alcoholic beverage, non-alcoholic beverage, dairy produce, fruit and vegetables, bakery, confectionery, ready meals, and other |
| Manufacturers and packers - non-food | Textiles and garments, electronics, car manufacturer, furniture, chemical plant, pharmaceuticals, printing, and engineering |
| Warehouses |  |
| Distributors and transporters | Wholesalers, haulage company, and food distributors |
| Retailers | Supermarket, small retailers and other |
| First responders | Ambulance, fire services, police |
| Military sites | Army, navy and air force |
| Restaurants and caterers | Restaurant/café/canteen, hotel/guest house, pub/club, take-away, mobile food unit, and other |
| Offices |  |
| Close contact services | Hairdressers, barber shops, beauty and nail bars, make-up studios, tattoo studios, tanning salons or booths, spas and wellness businesses, sports and massage therapy, wellbeing and holistic locations, dress fitters, tailors, and fashion designers |
| Other |  |

Outbreak sites were linked to workplaces in the NPD through their postcode and business name. If the outbreak site postcode and business name were incorrect, absent or not found in the NPD dataset, no match could be made and these SOI records were not included in the attack rate analysis. Furthermore, if the number of cases exceeded the number employed, the sites were excluded from the analysis. This may be due to underestimation of employment in the NPD for some workplace settings, such as crop production and warehouses where there is a reliance on temporary agency worker.

Geographical coordinates were added to HPZone and SOI data from the ONS Postcode Directory^15^, using the postcode of the outbreak settings.

## Results

Between 18 May and 12 October 2020, 1,317 confirmed workplace outbreaks were identified from HPZone, of which 1,305 could be mapped by postcode for the outbreak rate calculation. In addition, 390 outbreaks were identified from the SOI dataset, of which 285 could be linked directly to records in the NPD workplaces to add SIC and employment information. A further 21 outbreaks from the SOI dataset, where no case numbers were recorded or where the number of cases exceeded the number employed, were removed. This leaves 264 SOI records of outbreaks, including a total 2,649 confirmed COVID-19 cases, for the attack rate calculation.

The number of workplace outbreaks, including 1,317 outbreaks from HPZone and 390 outbreaks from SOI, by sector are listed in Table 2 and by region are listed in Table 3. The geographical distribution of the outbreaks sites that can be mapped by postcode to the NPD, including 1,305 outbreaks from HPZone and 285 outbreaks from SOI, are shown in Figure 1.

**Table 2.** The number of COVID-19 workplace outbreaks by sector (% the total) in England, May – Oct 2020

| **Workplace Setting** | **Count of Outbreaks (HPZone)** | **Count of Situations of Interest** |
| --- | --- | --- |
| Close contact services | 13 (1%) | 10 (3%) |
| Distributors and transporters | 84 (6%) | 19 (5%) |
| First responders | 48 (4%) | 16 (4%) |
| Manufacturers and packers of food | 117 (9%) | 96 (25%) |
| Manufacturers and packers of non-food | 195 (15%) | 41 (11%) |
| Military sites | 9 (1%) | 7 (2%) |
| Offices | 193 (15%) | 27 (7%) |
| Primary producers | 8 (<1%) | 7 (2%) |
| Restaurants and caterers | 53 (4%) | 26 (7%) |
| Retailers | 219 (17%) | 33 (8%) |
| Warehouses | 58 (4%) | 22 (6%) |
| Other | 266 (20%) | 46 (12%) |
| No setting type assigned | 54 (4%) | 40 (10%) |
| **Total** | **1,317 (100%)** | **390 (100%)** |

**Table 3.** The number of COVID-19 workplace outbreaks by region (% the total) in England, May – Oct 2020

| Region | Count of Outbreaks (HPZone) | Count of Situations of Interest |
| --- | --- | --- |
| East Midlands | 136 (10%) | 101 (26%) |
| East of England | 73 (6%) | 50 (13%) |
| London | 147 (11%) | 8 (2%) |
| North East | 39 (3%) | 22 (6%) |
| North West | 355 (27%) | 33 (8%) |
| South East | 62 (5%) | 18 (5%) |
| South West | 92 (7%) | 32 (8%) |
| West Midlands | 214 (16%) | 67 (17%) |
| Yorkshire and The Humber | 199 (15%) | 59 (15%) |
| Total | 1,317 (100%) | 390 (100%) |

**Figure 1.**
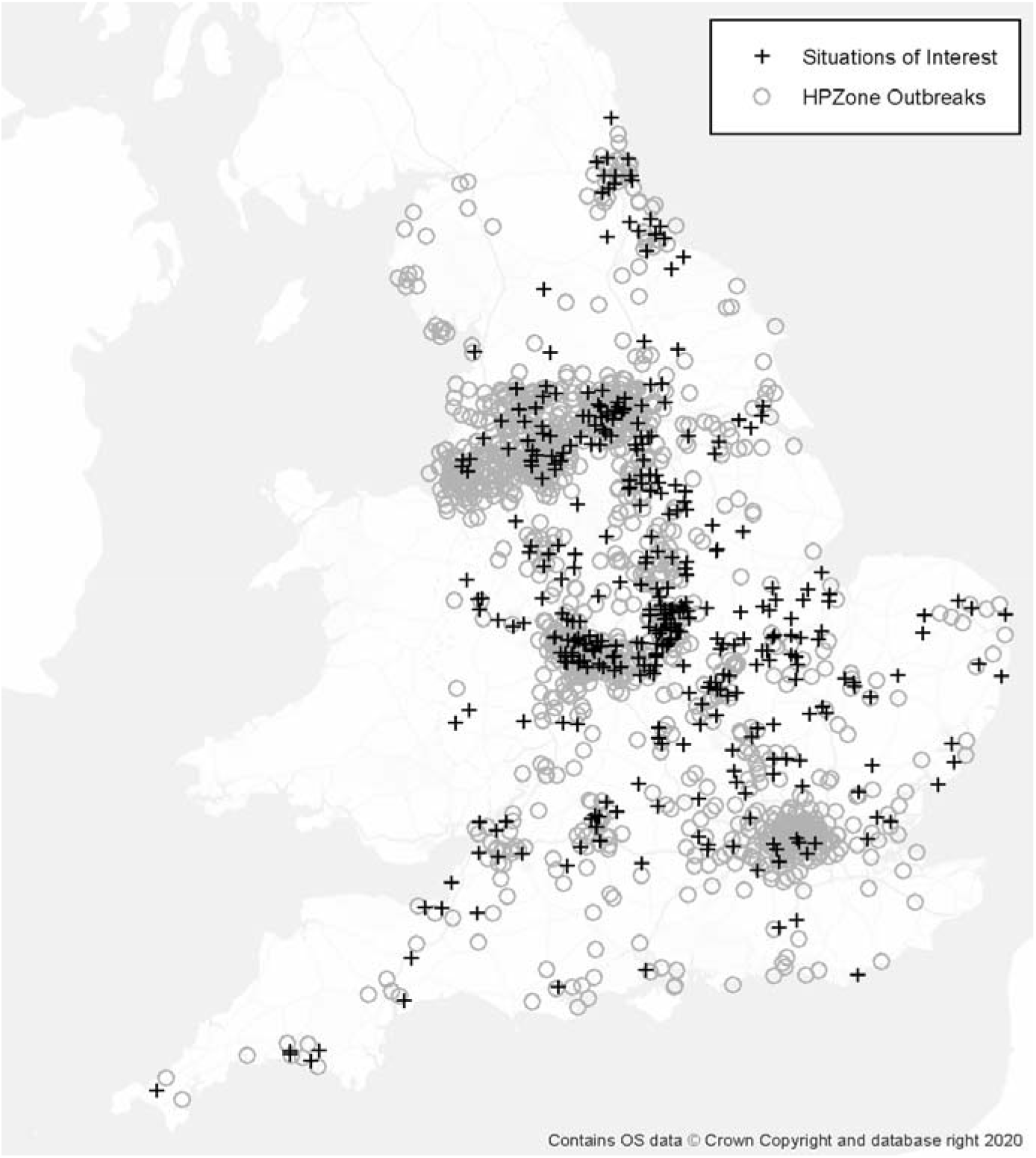
Geographical distributions of COVID-19 workplace outbreaks in England, May – Oct 2020

### Outbreak rates by geographical area (Region, UTLA)

Of the nine geographical regions in England, the North West of England had the highest number of outbreaks, affecting 351 workplaces, as well as the highest rate of outbreaks (155/100,000 workplaces) (Table 4). Of the 151 Upper Tier Local Authorities (UTLAs), the largest numbers of workplace outbreaks were mainly observed in northern English towns and cities with the highest outbreak rates registered in Blackburn with Darwen (387/100,000), Sandwell (351/100,000), Liverpool (349/100,000), Rochdale (277/100,000), Manchester (275/100,000) and Bradford (254/100,000) (Table 5).

**Table 4.** Number and rate of COVID-19 workplace outbreaks by English Region, May-Oct 2020

| Region | Number of Outbreaks | Number of Workplaces (England) | *Outbreak Rate* (per 100,000) |
| --- | --- | --- | --- |
| North West | 351 | 226,576 | 155 |
| Yorkshire and The Humber | 198 | 168,184 | 118 |
| West Midlands | 215 | 183,534 | 117 |
| East Midlands | 134 | 156,900 | 85 |
| North East | 39 | 67,056 | 58 |
| London | 149 | 375,249 | 40 |
| South West | 84 | 215,640 | 39 |
| East of England | 71 | 226,190 | 31 |
| South East | 64 | 349,945 | 18 |
| Total | 1,305 | 1,969,274 | 66 |

**Table 5.**
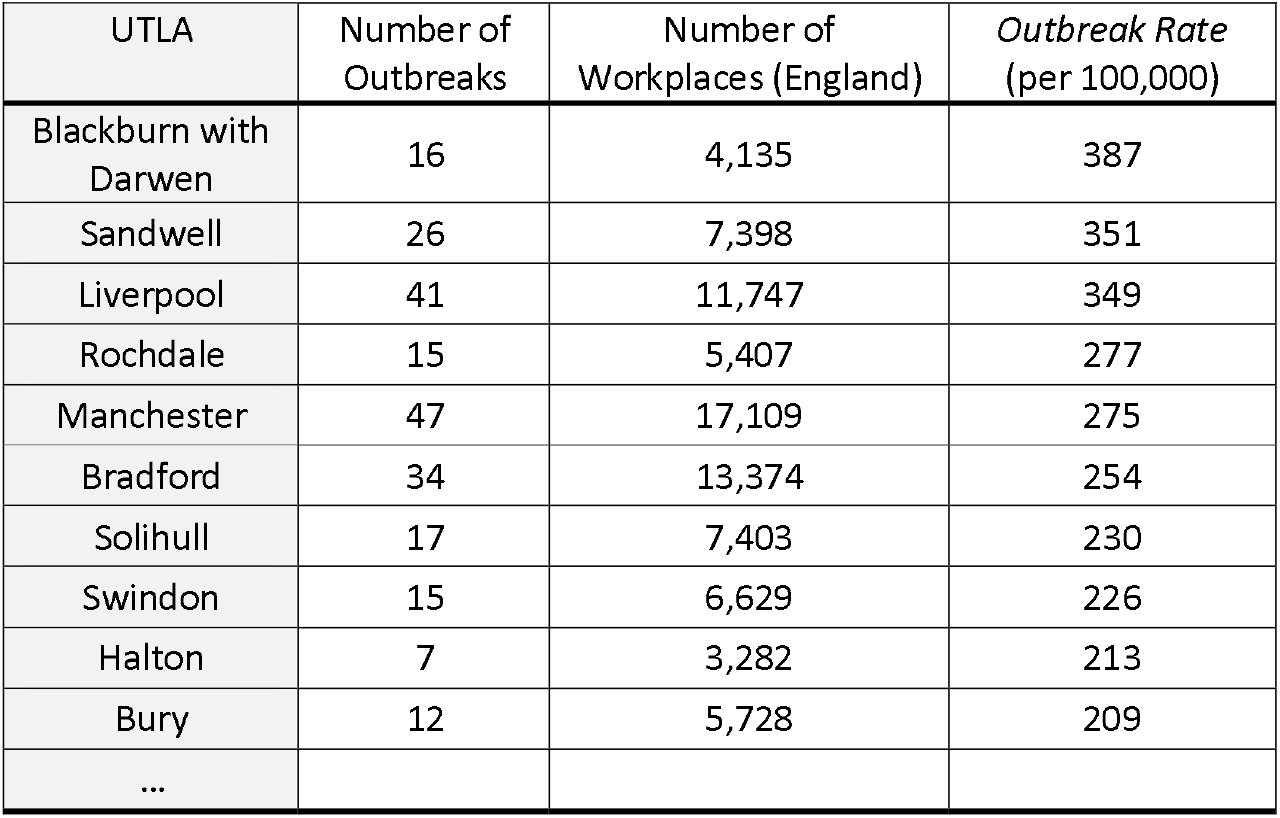
Number and the top 10 rates of COVID-19 workplace outbreaks in English Upper Tier Local Authority (UTLA), May-Oct 2020

### Outbreak rates by workplace setting

In comparison with other sectors, retailers had the highest number of outbreaks, affecting 219 workplaces, followed by manufacturers and packers of non-food products (195) and offices (193). However, after applying the denominator data, the highest outbreak rate was in manufacturers and packers of food (1,672/100,000), based on 117 outbreaks out of 6,998 workplaces. This was much higher than the outbreak rates for the remaining sectors with warehouses and manufacturers and packers of non-food products the next highest at 385 per 100,000 workplaces and 308 per 100,000 workplaces respectively (Table 6).

**Table 6.** Number and rate of workplace outbreaks by sector in England, May-Oct 2020

| Workplace Setting Type (from HPZone) | Number of Outbreaks | Number of Workplaces (England) | *Outbreak Rate* (per 100,000) |
| --- | --- | --- | --- |
| Manufacturers and packers of food | 117 | 6,998 | 1,672 |
| Warehouses | 58 | 15,058 | 385 |
| Manufacturers and packers of non-food | 195 | 63,312 | 308 |
| Retailers | 219 | 195,025 | 112 |
| First responders/Military sites | 57 | 67,257 | 85 |
| Distributors and transporters | 84 | 125,414 | 67 |
| Restaurants and caterers | 53 | 117,836 | 45 |
| Offices | 193 | 721,351 | 27 |
| Close contact services | 13 | 52,866 | 25 |
| No setting type assigned | 54 | 511,071 | 11 |
| Primary producers | 8 | 93,086 | 9 |
| Other | 266 | - | - |
| Total | 1,317 | 1,969,274 | 67 |

### Outbreak rates by region and workplace setting type

High outbreak rates in manufacturers and packers of food were observed consistently across seven English regions, including: the West Midlands (3,555/100,000 workplaces), Yorkshire and the Humber (3,132/100,000 workplaces), the North West (2,926/100,000 workplaces), the East Midlands (2,031/100,000 workplace), the East of England (1,664/100,000), the North East (1,282/100,000 workplaces), and South West (638/100,000 workplaces) (Table 7). In addition, high rates of outbreaks were observed in warehouse settings in the East Midlands and the North West with an outbreak rate of 1,524 per 100,000 workplaces and 793 per 100,000 workplaces respectively (Table 7).

**Table 7.** The top 10 outbreak rates by English region and sector combined, May-Oct 2020

| Region – Workplace Setting Type (from HPZone) | Number of Outbreaks | Number of Workplaces | *Outbreak Rate* (per 100,000) |
| --- | --- | --- | --- |
| West Midlands – Manufacturers and packers of food | 23 | 647 | 3,555 |
| Yorkshire and The Humber - Manufacturers and packers of food | 28 | 894 | 3,132 |
| North West – Manufacturers and packers of food | 28 | 957 | 2,926 |
| East Midlands – Manufacturers and packers of food | 13 | 640 | 2,031 |
| East of England – Manufacturers and packers of food | 12 | 721 | 1,664 |
| East Midlands - Warehouses | 19 | 1,247 | 1,524 |
| North East – Manufacturers and packers of food | 4 | 312 | 1,282 |
| North West – Manufacturers and packers of non-food | 65 | 8,074 | 805 |
| North West - Warehouses | 15 | 1,891 | 793 |
| South West – Manufacturer and packers of food | 6 | 940 | 638 |
| ... |  |  |  |

### Attack rates by enterprise size

A minority (29%) of the outbreaks recorded in SOI were in small enterprises (<50 employees) but the proportion of small enterprises was higher for close contact services (83%) and restaurants and caterers (56%) (Table 8). The overall median attack rate was 3.4% for outbreaks in all enterprises. The attack rates increased as the number employed at a workplace decreased (Table 9).

**Table 8.** Number and proportion of the workplace outbreaks in the small enterprises (<50 employees), in England, May-Oct 2020

| Workplace Setting Type | Outbreaks at workplaces with <50 employees | Total outbreaks | Proportion at workplaces with <50 employees |
| --- | --- | --- | --- |
| Close contact services | 5 | 6 | 83% |
| Distributors and transporters | 1 | 16 | 6% |
| First Responders/Military Sites | 7 | 19 | 37% |
| Manufacturers and packers of food | 11 | 82 | 13% |
| Manufacturers and packers of non-food | 8 | 29 | 28% |
| Offices | 9 | 25 | 36% |
| Other | 8 | 25 | 32% |
| Primary producers | 2 | 5 | 40% |
| Restaurants and caterers | 9 | 16 | 56% |
| Retailers | 13 | 28 | 46% |
| Warehouses | 1 | 13 | 8% |
| No setting type assigned | 10 | 21 | 48% |
| Total | 84 | 285 | 29% |

**Table 9.** Median and IQR attack rates of workplace outbreaks by enterprise size (number of employees) in England, May-Oct 2020

| Enterprise size | Individual Outbreaks |  |  |  | Workplaces |  | Attack Rate |  |
| --- | --- | --- | --- | --- | --- | --- | --- | --- |
|  | Count of cases | Count of sites | Cases per site |  | Number employed (NPD) at outbreak sites |  | Cases per 100 employed |  |
|  |  |  | Median | IQR | Median | IQR | Median | IQR |
| 0-49 employees | 290 | 68 | 3 | 2 | 19 | 17 | 17.8 | 20.5 |
| 50-249 employees | 622 | 82 | 5 | 6 | 119 | 90 | 4.3 | 6.4 |
| 250 plus employees | 1,737 | 114 | 6 | 9 | 622 | 587 | 1.1 | 1.3 |
| Total | 2,649 | 264 | 4 | 6 | 176 | 473 | 3.4 | 11.3 |

### Attack rates by workplace setting

Outbreaks in close contact services had the largest attack rate (median 16.5%), which was based on 22 test-confirmed cases at 6 outbreak sites (Table 10). The attack rates were also high for outbreaks in restaurants and caterers (median 10.3%), based on 49 test-confirmed cases at 14 sites; and in manufacturers and packers of non-food products (median 6.7%), which was based on 270 cases at 29 sites.

**Table 10.**
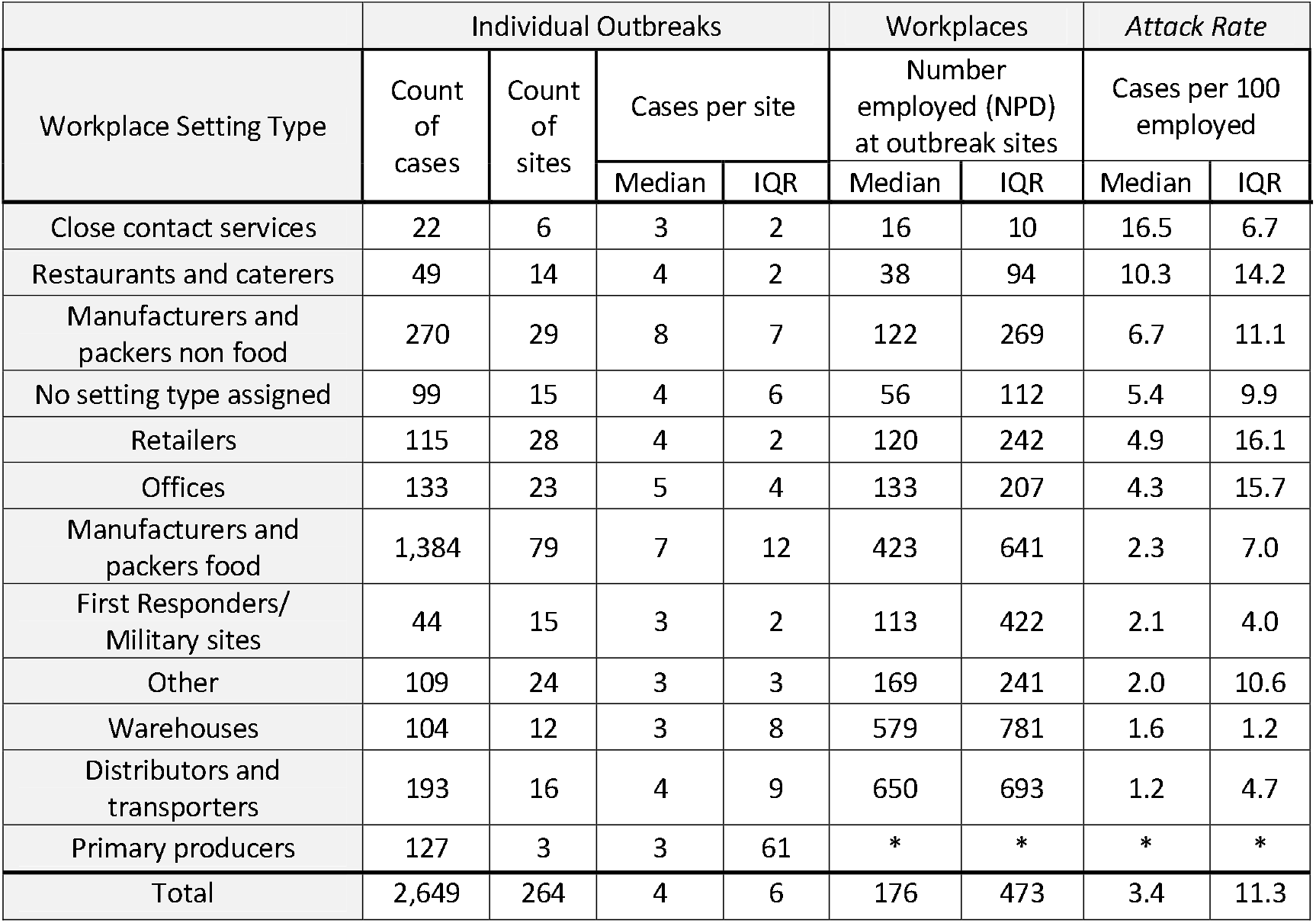
Median attack rates of workplace outbreaks by sector in England, May-Oct 2020 Cases by Setting Type for settings (including all enterprises)

The distribution of the number of workplace outbreaks (a total of 264) by attack rate is shown in Figure 2. Most of the outbreaks (162 outbreaks) had an attack rate less than 6%. However, in a small number of outbreaks (57) the attack rate was over 15%. The geographical distribution of the outbreaks by attack rate is shown in Figure 3.

**Figure 2.**
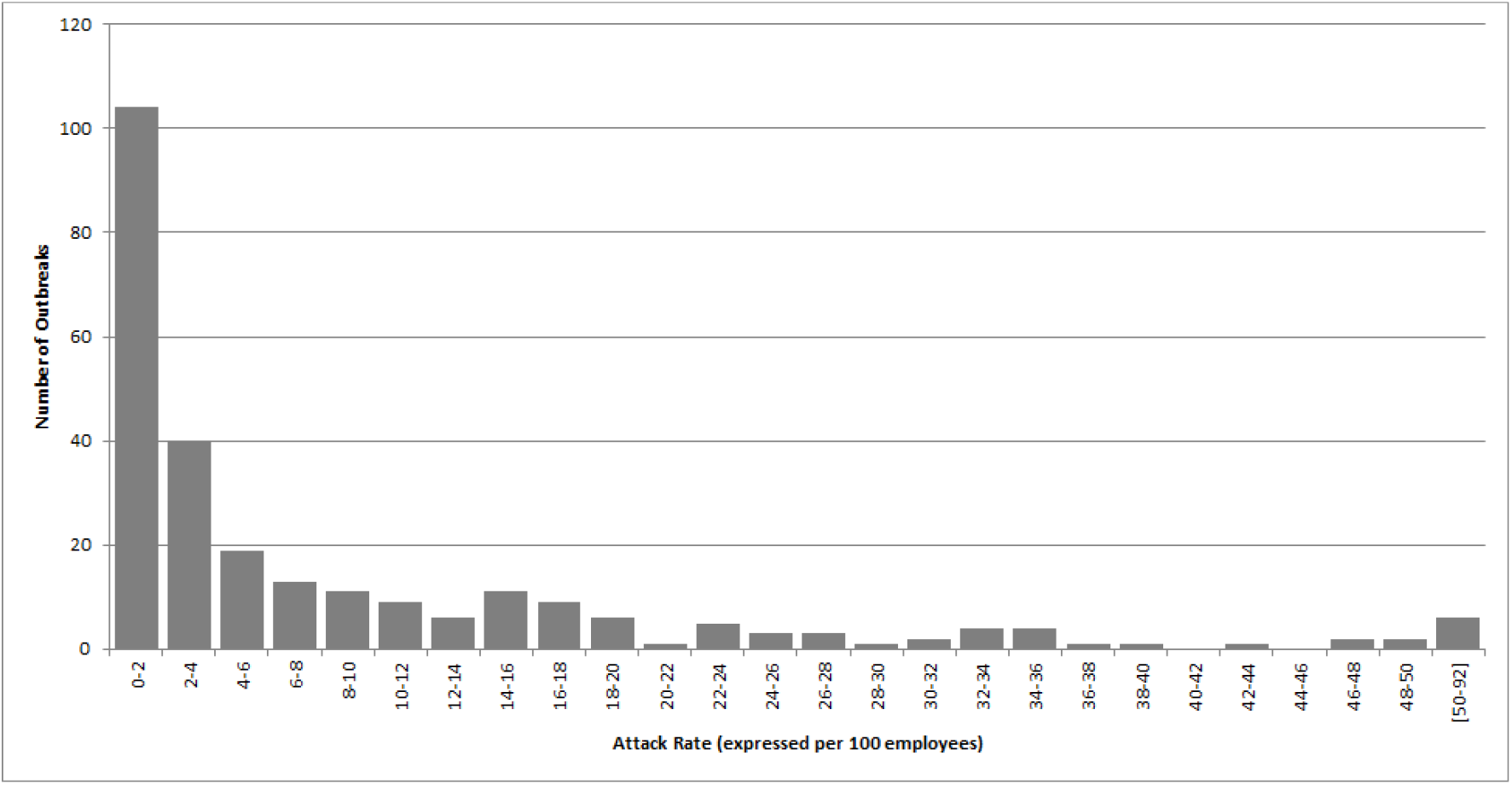
Distribution of the number of outbreaks by attack rate in England, May-Oct 2020

**Figure 3.**
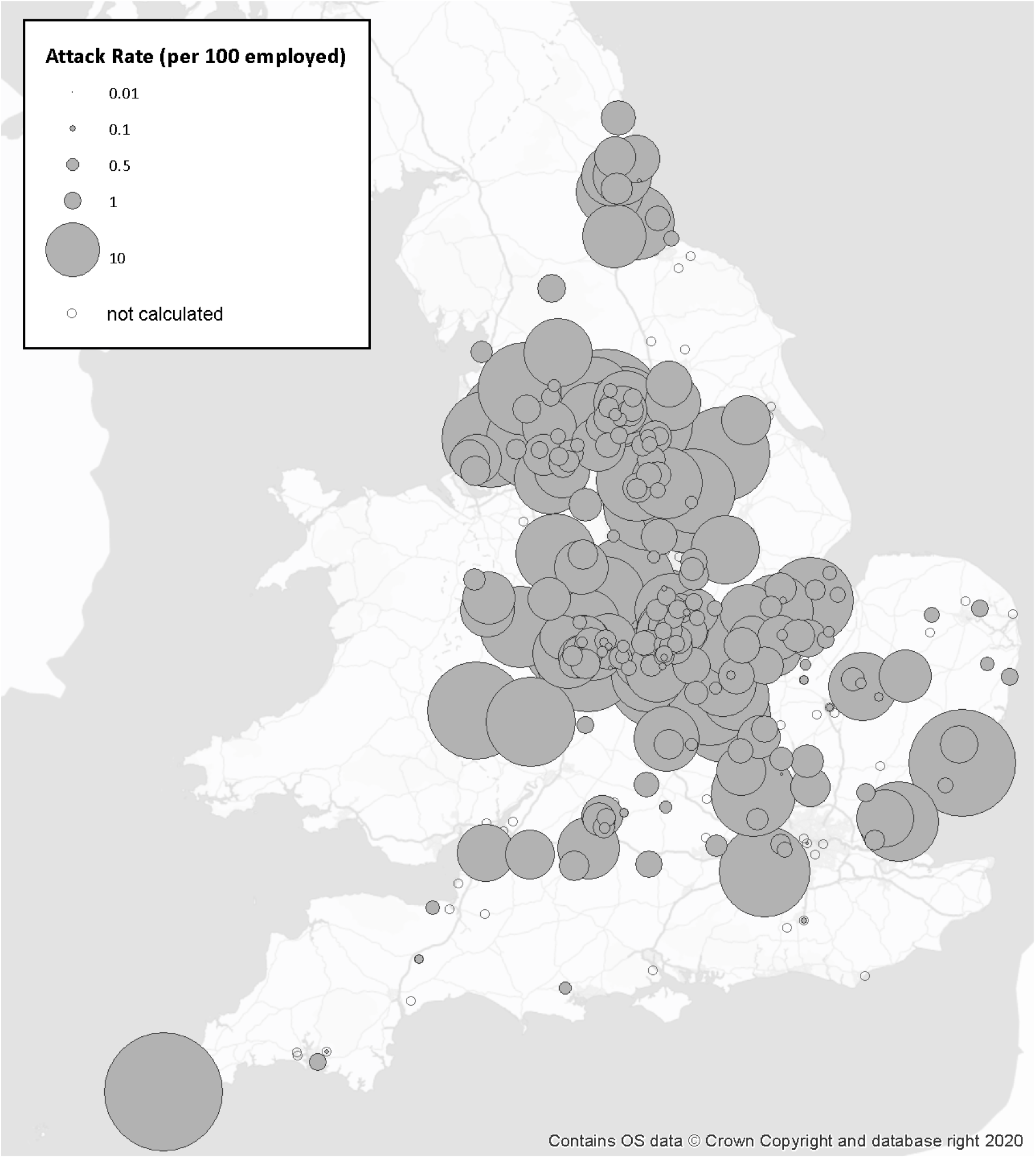
Geographical distributions of COVID-19 outbreaks by *Attack Rate* in England, May – Oct 2020

## Discussion

Our study has, for the first time, utilised the number of confirmed COVID-19 outbreaks recorded in PHE information system and combined them with relevant denominator data held by HSE to calculate outbreak rates and attack rates by sector and geographical area. A relatively large number of outbreaks were observed in some workplace settings, including retail, manufacturers and packers of non-food products and offices. When we applied the denominator data of the total number of the relevant settings, manufacturers and packers of food had the highest outbreak rates and this was consistent across seven English regions. These findings have provided some indication of the relative risk of outbreaks in different workplace settings. Manufacturers and packers of food are part of the national infrastructure and these workplaces were kept open throughout the pandemic even during the national lockdown. Outbreaks of COVID-19 in manufacturers and packers of food have been frequently reported in the literature and in the media in many counties^16^. However, only a few studies have investigated the potential transmission risk factors in this type of workplace settings ^17^. In addition, it will be important to continue to monitor outbreak rates in different workplace settings and by industrial sector as the country is moving out of the pandemic and more sectors are increasing their work capacity.

Our study has also, for the first time, utilised data from the public health COVID-19 outbreak management records to calculate infection attack rates. This allows comparison of the potential extent of transmission between outbreaks in different workplace settings. Close contact services and restaurants/caterers had the highest attack rates which were mostly associated with outbreaks in small enterprises. Manufacturers and packers of non-food products also had relatively large attack rates but were mostly associated with outbreaks in medium and large enterprises. A distribution of the number of workplace outbreaks by attack rate has been generated. However, it is worth noting that the SOI dataset are skewed towards large and more impactful outbreaks. Furthermore, more detailed analysis of outbreak rates is limited by low numbers of outbreaks in certain industrial sectors, such as primary producers which include fruit and vegetable growers, animal and animal products.

Our analysis carried some limitations. The potential under-identification of outbreaks in small enterprises (<50 employees) in the numerator coupled with the vast number of small enterprises in the denominator may greatly underestimate the outbreak rates. This could particularly impact on small business-dominated sectors, such as close contact services and restaurants/caterers, where estimated outbreak rates were relatively low, but attack rates were relatively high.

The number of outbreaks reported to HPZone could be affected by national and local level operational changes. For example, as caseload increased in September and October 2020, some HPTs transferred the management of some outbreaks/clusters to local authorities. As a result, HPZone no longer represents a comprehensive list of COVID-19 outbreaks/clusters in England. The impact of this change is unclear. This practice also varies regionally, which affects the ability to measure the changes of outbreak occurrence or outbreak rate over time, as well as the ability to measure regional variations using HPZone data.

SOI outbreak data is a subset of the HPZone outbreaks/clusters with no clear selection criteria. Data entry was through a separate mechanism and was an additional task for HPTs, with no perceived added benefit to the operational management of the outbreaks/clusters. The proportion of HPZone outbreaks/clusters in the workplace being reported as SOI decreased over time, especially from September 2020 onward as HPTs were under pressure to respond to an increasing number of outbreaks. This means the SOI data are not representative of all outbreaks responded to by HPTs in England.

NPD workplaces information also has some limitations in providing reliable working population data as the denominator, which may cause imprecisions in the attack rate calculation. Workplaces vary in size from single person enterprises with no further employees to major sites, employing thousands of workers. Workplaces may also close, expand and diversify over relatively short time scales due to commercial or financial pressures. In our study, NPD data represent the distribution of the GB population pre-pandemic; the number of employees in some workplace settings will be reduced due to social distancing measures. This may cause underestimation of the attack rates due to overestimation of the denominator. The level of underestimation varies by sector with some sectors completely closed and others kept operating in full capacity throughout the pandemic. However, the impact of this limitation may attenuate as society gradually opens.

In addition, the NPD workplaces information may not capture the number of employees in the transient workforce or working in irregular patterns. For example, employees on temporary contracts or seasonal workers in the agriculture sector might not be accounted for in the NPD. Employees in some other workplaces, such as in distribution centres, transportation of goods between depots, and in construction will be accounted for but their non-fixed working locations will not be well-represented by a single geographical reference (e.g. postcode of the company address). Similarly, agency staff and sub-contractors are unlikely to be accounted for at the location where they carry out their work activities. This may cause over estimation of the attack rate due to the underestimation of the denominator. In addition, each workplace in the denominator data is classified to a single sector, such as manufacture of food products (SIC 10); secondary settings such as offices within a workplace could not be captured. This will affect the rate calculation for these less well-defined setting types such as offices and warehouses.

The information on the number of employees in a company is extracted from the Office for National Statistics (ONS) Inter-Departmental Business Register (IDBR)^13^. In the IDBR, some enterprises are located to a PO Box which can be situated at Royal Mail distribution centre; this accounts for 1.8% of employment in the NPD workplaces layer. A large organisation that operates through different locations may be represented by a single record situated at the company headquarters. Consequently, the employee number at the headquarters might be over-estimated while the employee numbers at the secondary locations of the company may not be represented.

Early identification of COVID-19 outbreaks/clusters and visualisation of their geographical distribution can provide a rapid assessment of where the SARS-CoV-2 transmission is occurring. A large number of COVID-19 outbreaks/clusters have been reported, both in scientific literature and in the media, in a wide range of mostly indoor settings across the world ^3 18^. Most of the COVID-19 clusters will be in residential settings, particularly in households, due to the increased risk of transmission caused by close and frequent contact. The COVID-19 surveillance report published by PHE on 11^th^ February 2021 has shown that over 90% of the confirmed cases, according to their type of residence in week 2-5 in 2021, were in residential dwelling, including houses, flats, and sheltered accommodation^19^. However, a household cluster will not result in a large outbreak without the virus spreading beyond the household setting. Some of these individuals in households could also travel to other settings including the workplaces. Transmission is a continuous risk. It is difficult to establish where transmission really occurred. Community transmission will also occur through social gathering, particularly gathering outdoors, shopping in supermarkets or using public transport. However, it is difficult to identify outbreaks/clusters from the large number of transient populations in these settings without a rigorous surveillance system for widespread testing and detailed contract tracing. This may underestimate the relative importance of the potential transmission in these less well-defined settings or population.

Since our study, the approach of utilising the suitable denominator data to calculate outbreak rates has been adopted by the Joint Bio-security Centre (JBC) and will be embedded in their regular national surveillance analysis and reporting on workplace outbreaks and outbreaks rates. Although our study was only able to analyse the workplace outbreak data, the same approach can be applied to the calculation of outbreak rates and attack rates in other geographical locations and other types of settings, such as care homes, hospitals, schools and prisons. These will potentially guide interventions to target high risk areas and to limit the spreading of the virus.

Our study was not able to assess the potential changes in COVID-19 outbreak rates and attack rates over time due to, in part, the limited time period of data (May-Oct 2020) and the inconsistency in recording outbreaks/clusters in the HPZone and SOI datasets. Further consideration will be to analyse the more enhanced outbreak/cluster data collected from NHS Test and Trace over a longer period of time of this pandemic to identify past and emerging trends.

Evidence shows that there could be marked heterogeneity in the characteristics of SARS-CoV-2 transmission^4^, with the majority (∼80%) of the secondary transmission caused by a very small proportion of SARS-CoV-2 infected persons, and outbreaks of COVID-19 distributed unevenly in certain settings and geographical locations^20^. Our study has found increased rates of outbreak in certain industrial sectors and English regions, and a large variation of the size of the attack rates. The variation of the rates may be impacted by the type of work activities, the size of the enterprises, the transmission risk and the intervention strategies to limit the transmission in these sectors. The risk of transmission will also be associated with the behavioural and social factors of the individuals, the environment and the control measures that influence transmission dynamics of the virus in certain settings^3^.

The current study has investigated the patterns and rates of COVID-19 outbreaks in England. Further studies, as part of the National Core Study programme, will investigate and identify the characteristics of the outbreak settings that could increase risk of transmission. We have also designed and commissioned comprehensive epidemiological field studies to collect data from live COVID-19 outbreaks in workplace settings in order to better understand the transmission risk factors and transmission routes^8^.

## Data Availability

The data used to support the findings of the study are included in the references within this paper.

## Author roles

Conceptualization, Y.C., T.A., and F-M.K.; Methodology, T.A., Y.C., and C.F.F.; Validation, Y.C., T.A., C.F.F., and F-M.K.; Formal Analysis, T.A.; Investigation, Y.C., T.A., and C.F.F.; Data Curation, T.A. and C.F.F.; Writing-Original Draft Preparation, Y.C., T.A., and C.F.F.; Writing-Review & Editing, Y.C., T.A., C.F.F., and F-M.K.; Visualization, T.A.; Supervision, Y.C. and F-M.K.; Project Administration, F-M.K. and Y.C.; Funding Acquisition, UK COVID-19 National Core Studies Consortium.

## Disclaimer

The opinions and assertions contained herein are private views of the authors and do not necessarily reflect those of the Health and Safety Executive or Public Health England.

## Copyright

© Crown copyright (2021), Health and Safety Executive. This is an open access article distributed under the terms of the Open Government Licence v3.0, which permits re-use, distribution, reproduction and adaptation, provided the original work is properly cited.

